# Evidence for a protein leverage effect on food intake in a Norwegian population

**DOI:** 10.1101/2025.11.07.25339754

**Authors:** Rikke Eriksen, Kamilla Rognmo, Laila A Hopstock, James E McCutcheon

**Author notes:** Contact information: James E McCutcheon, Dept. of Psychology, UiT The Arctic University of Norway, Huginbakken 32, 9019 Tromsø, Norway.

## Abstract

In this pre-registered study, we aimed to explore the protein leverage hypothesis in a general population, by studying the relationship between habitual dietary protein intake, total energy intake, and body mass index (BMI), and whether proportion of ultra-processed food (UPF) was associated with any of these variables. We used regression-based analyses to investigate these phenomena in cross-sectional data from a Norwegian population-based study, the seventh survey of Tromsø Study 2015-2016, (*n* = 11,152; 40-99 years). Total energy intake was negatively associated with proportion of dietary protein (*L* = -0.36, *p* < .001) and positively associated with dietary fat (*L* = 0.33, *p* < .001). Although we planned to test the relationship between BMI and dietary protein, there was no positive association between total energy intake and BMI meaning that these data were not suitable for testing an effect of protein leverage on BMI. Proportion of UPFs was positively associated with total energy intake (*b* = 554, *p* < .001), negatively associated with proportion of dietary protein (*b* = -2.0, *p* < .001), and positively associated with BMI (*b* = 0.011, *p* = .026). In summary, our study of middle-to-older aged Norwegians provides strong support for a protein leverage effect on energy intake.

## 1. Introduction

The obesity epidemic is a global problem that is estimated to contribute to a huge number of deaths each year as well as posing a significant economic burden (World Health Organization, 2021). Although its causes are multifactorial, reduced intake of dietary protein has been implicated via the protein leverage hypothesis. The protein leverage hypothesis is based on a nutritional geometric framework that was developed to explain how humans and other animals meet their nutritional needs (Raubenheimer et al., 2022). The hypothesis emphasizes that protein is the most essential macronutrient as many of the amino acids cannot be synthesized de novo and must be acquired from one’s diet. This implies that humans need to consume a target level of protein on a regular basis (i.e., on the order of days) resulting in a strong appetite for protein (Raubenheimer et al., 2015). The protein leverage mechanism is proposed to regulate energy intake towards this protein target through appetite (Simpson & Raubenheimer, 2005) and, as such, the proportion of protein in a given diet is suggested to influence food intake more strongly than total calories consumed. Accordingly, if a food environment is “protein diluted”, (i.e., contains a lower proportion of dietary protein than a balanced diet), excess energy will be consumed in an effort to reach the protein target (Simpson & Raubenheimer, 2005). Recent analyses find that the US food supply has indeed become protein diluted over recent decades, and though this dilution is small (∼1% of available calories), modeling suggests a significant effect on development of obesity in the adult population (Hall, 2019).

Several studies, including randomized controlled trials (Gosby et al., 2011; Martens et al., 2013) and observational studies (Honfo et al., 2023; Saner et al., 2023; Saner et al., 2020; Zhang et al., 2025; Zhang et al., 2023), find support for the energy regulating effects of the protein leverage mechanism in specific cohorts including preschool children prone to obesity, children and adolescents living with obesity, and older adults. To our knowledge, few observational studies have examined protein leverage effects in adults and only one (Zhang et al., 2025) has been conducted in European populations.

The dilution of protein in the human food supply (Raubenheimer & Simpson, 2023) may be linked to the increase in availability and popularity of ultra-processed foods (UPFs) . The human obesity epidemic has developed concomitantly with the emergence of these foods, and now UPFs dominate the market in several high-income countries. For example, in the UK, US, and Canada more than half of the energy consumed comes from these foods (Monteiro et al., 2019). In the context of protein leverage, high UPF diets that tend to be low in dietary protein should lead to an increase in total calories consumed. Indeed, this has been found in national representative samples of the Australian (Grech et al., 2022a) and US (Martínez Steele et al., 2018) populations, which show that high UPF diets are related to a lower proportion of dietary protein intake and a heightened energy intake. In addition, a carefully planned 14-day RCT (Hall et al., 2019) found overcompensation of energy intake on a UPF diet (∼16% protein) when compared to an non-UPF diet (∼19% protein). This increased energy intake was accompanied by an increase in bodyweight where participants gained 0.9 kg when on the UPF diet and lost 0.9 kg when on the non-UPF diet, implying that weight gain observed in high UPF diets could result from unbalanced protein levels. In fact, replotting of data from Hall et al. (2019) in Raubenheimer and Simpson (2023) suggests that the protein leverage mechanism accounts for up to 100% of the observed change in energy intake between the UPF and non-UPF diets. Similar results were found in an RCT where participants gained more weight and increased energy intake on a UPF diet than on a non-UPF diet, while no significant differences were found in protein intake between the diets (Hamano et al., 2024).

The biological processes that underpin protein leverage are still under study, but one potential mechanism is variation in levels of the hormone fibroblast growth factor 21 (FGF21). Studies in humans and rodents have found that FGF21 increases in response to protein restriction (Laeger et al., 2014; Volcko & McCutcheon, 2022). In mice, FGF21 has also been found to increase protein intake (Solon-Biet et al., 2023) and protein preference without increasing total food intake (Hill et al., 2019). In humans, plasma FGF21 levels have been found to be negatively associated with percent protein in the diet (Gosby et al., 2016; Nicolaisen et al., 2025). FGF21 increases manyfold in response to a low-protein overfeeding diet (Hollstein et al., 2019), indicating that its release is not just dependent on the caloric content of the food. Additionally, FGF21 levels after exposure to a low-protein overfeeding diet are positively related to 24 hour energy expenditure (Vinales et al., 2019). Similarly, energy requirements increased concomitantly with FGF21 levels in lean men on a protein restricted diet (Nicolaisen et al., 2025). As such, the relationship between FGF21, protein intake, and bodyweight is complex.

In our study, we aimed to replicate and extend findings on the protein leverage mechanism by testing the predicted relationships between dietary protein, energy intake, and body mass index (BMI) in middle-aged and older adults through two preregistered hypotheses (osf.io/4g62f). These were that (1) the proportion of dietary protein will be negatively associated with total energy intake, and (2) the proportion of dietary protein will be negatively associated with BMI. In addition, we assessed properties of diets high in UPFs. Finally, we investigated the relationship between habitual dietary protein intake and FGF21.

## 2. Methods

### 2.1 Data source and sample

The current study includes data from the Tromsø Study, a large-scale population-based study conducted in the municipality of Tromsø, Norway. To date, seven surveys have been carried out and the eight survey is ongoing (Tromsø1–Tromsø8, 1974–2026). Data collection includes questionnaires and interviews, biological sampling and clinical measurements. This analysis includes data from Tromsø7 (2015–2016), where all inhabitants of Tromsø municipality aged 40 years and older (*N* = 32,591; 50.7 % women) were invited, and 21,083 (65%; 52.5 % women) attended. Details of the study design, sample, and data collection methods for Tromsø7 have been described elsewhere (Hopstock et al., 2022).

The current study was approved by the Regional Committee for Medical Research Ethics Northern Norway (ref. 789456, 14/11/2024), and the Norwegian Agency for Shared Services in Education and Research (ref. 935030, 9/10/2024). All participants gave written consent at study attendance. Data from participants that later withdrew their consent were not used in the analysis. The current study analyzed data from 11,152 participants. The exclusion criteria are visualized in the flow chart depicted in Figure 1. Data exclusion plan and two hypotheses were preregistered on Open Science Framework (osf.io/4g62f).

**Figure 1.**
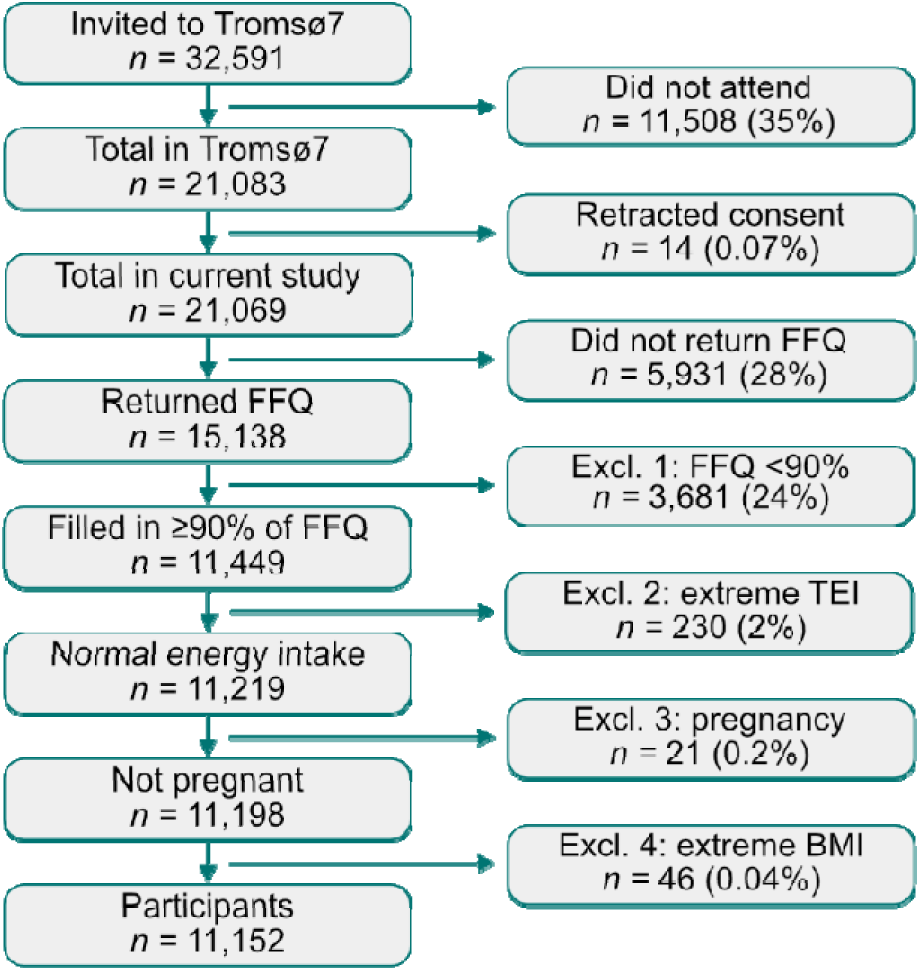
Flowchart of exclusions leading to final sample. Tromsø7 data was collected in 2015-2016. Food frequency questionnaire (FFQ). % indicates percent change since the last step. To increase the validity of the data we excluded (1) participants that completed less than 90% of the FFQ and (2) participants with extreme intakes of energy, i.e., the 1^st^ (below 3,950 kJ/day) and 99^th^ (above 21,249 kJ/day) percentile in total energy intake (TEI). Pregnancy may interfere with habitual eating patterns and body composition and we therefore excluded (3) participants that reported being pregnant at the time of the data collection. Finally, we excluded (4) participants with extreme BMI values, i.e., groups of BMI with less than 10 participants, ensured by binning the BMI data in bins of one BMI unit and excluding bins with fewer than 10 observations.

### 2.2 Food Frequency Questionnaire

All participants were invited to complete a food frequency questionnaire (FFQ) designed to collect data on habitual dietary patterns by assessing frequency and quantity of intake of 261 of single foods, beverages, dishes and dietary supplements (Carlsen et al., 2010). Details of the FFQ used in Tromsø7 have been described elsewhere (Lundblad et al., 2019). The FFQ was developed and validated (Carlsen et al., 2010) at the University of Oslo, and is designed to detect dietary habits in Norwegian adults. Food, energy and nutrient intakes were calculated using the food database KBS AE14 and KBS software system (KBS, version 7.3.), based on the Norwegian food composition tables from 2014 to 2015.

### 2.3 Demographics and Health Related Information

Height and weight were measured by trained personnel using a Jenix DS-102 scale (DongSahn Jenix, Seoul, Korea). BMI was calculated as kg/m^2^ and categorized in the standard groups normal (<25 kg/m^2^), overweight (25–<30 kg/m^2^), and obese (≥30 kg/m^2^). The group normal includes a low proportion of underweight participants (≤1 %) as reported in Løvsletten et al. (2020).

Demographic and health information was collected by questionnaires. Level of leisure time physical activity was measured with the Saltin-Grimby Physical Activity Level Scale (Grimby et al., 2015) including sedentary, light, moderate, and vigorous physical activity levels. Education level included primary/partly secondary education (≤10 years), upper secondary (10 + minimum 3 years), tertiary short (college/university less than 4 years), and tertiary long (college/university 4 years or more). Current or previous disease included the four main non-communicable diseases (NCD) from the NCD framework (World Health Organization, 2013): cardiovascular diseases (myocardial infarction and/or stroke), cancer, chronic respiratory diseases (asthma and/or chronic bronchitis/emphysema/chronic obstructive lung disease) and/or diabetes, in addition to psychiatric problems the participant reported to have sought help for. Presence of chronic pain was defined by the question ”Do you have persistent or constantly recurring pain that has lasted for 3 months or more?” with answer options “yes” or “no”. Smoking status was defined by the question “Do you/did you smoke daily” with answer options “yes, now”, “yes, previously”, or “never”.

### 2.4 Fibroblast Growth Factor 21

Non-fasting blood samples were obtained with a light tourniquet released after venipuncture (Hopstock et al., 2022). In a randomly selected subsample (*n* = 1,144), FGF21 was analyzed as part of the OLINK Target Inflammation panel which uses Proximity Extension Assay (PEA) technology to measure relative protein levels.

### 2.5 NOVA Classification of Food Processing Level

The food items from the FFQ were categorized based on the NOVA classification system (Monteiro, 2009) and the procedure was adapted from Huybrechts et al. (2022) (Huybrechts et al., 2022). The NOVA system classifies foods and dishes into four mutually exclusive groups (Monteiro, 2009). Based on the categorization of the FFQ, a binary variable was made for (0) non-ultra-processed foods (NOVA groups 1-3) and (1) ultra-processed foods (NOVA group 4). For further details, see Supplementary Materials section 1.

### 2.6 Statistical Analyses

Data pre-processing and statistical analyses were performed using RStudio version 4.2.2. Data visualizations were made in Python version 3.9.

The protein leverage mechanism was tested through a power function (equation 1.1) and indexed by the L-value,

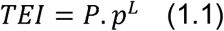

where TEI = total energy intake per day, *P* = a constant obtained from the analysis, *p* = the proportion of protein in the diet, and *L* = the strength of leverage. An *L*-value of -1 indicates complete protein leverage, while an *L-*value of 0 indicates that protein has no control over total energy intake (Senior et al., 2022). A positive *L*-value indicates that protein increases energy intake, i.e., the opposite of the predicted pattern. An *L*-value between -1 and <0 indicates partial protein leverage. The variables TEI and *p* are taken from the data set, while *P* and *L* are derived from the model when Equation 1.1 is log-transformed to a linear regression equation (equation 1.2),

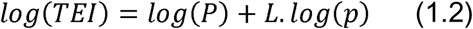

To assess the individual contributions of each macronutrient to total energy intake, energy density, and food dry weight, mixture models were run with the R package, *Mixexp* (Lawson & Willden, 2016). The package demands that the predictors (i.e., the three macronutrients) sum to one, which was accomplished by removing the contribution of alcohol and fiber to total energy intake. The mixture models were run based on the framework described by Lawson and Willden (2016), starting with a linear model and increasing the polynomial complexity (for details see Supplementary Materials section 2). The output from the mixture model was used to make mixture triangle surface plots, which plotted predicted outcome variables (total energy intake, energy density, and food dry weight) per participant based on macronutrient composition.

The proportion of UPFs in the diet was calculated as intake of UPFs in kJ/day divided by total intake of food in kJ/day (i.e., UPF proportion (%E) = UPF kJ /day/ total intake of food kJ/day). Analyses were conducted on UPF intake classified into quintiles, and for the effect of UPF intake on protein leverage, UPF intake was categorized as low or high via a median split. For relevant analyses, significant effects were followed up with contrast analyses using the *emmeans* package in RStudio.

Multiple linear regression models with relevant covariates were run with BMI as the outcome. This includes the following binary variables, which were entered into the model in the given order: age (over vs. under 60 years), sex (female vs. male), physical activity level (sedentary vs. three active levels), health conditions (presence vs. absence of conditions), education level (lowest vs. three highest levels), and smoking status (not current vs. current smokers). BMI was positively skewed and was therefore log-transformed prior to analyses.

For mixture modeling and BMI analyses, the Akaike Information Criterion (AIC) (Akaike, 1992) was used to choose between models, and if several models were within two AIC points of each other, the simplest model was favored.

### 2.7 Code and data availability

The data that support the findings of this study are available upon application to the Tromsø Study. Restrictions apply to the availability of these data, which were used under license for this study. All analysis code is available at https://github.com/jaimemcc/proteinleverage.

## 3. Results

### 3.1 Descriptive Statistics

Descriptive statistics for the full sample (*n* = 11,152, 53.3% women) are presented in Table 1. Information about BMI was missing for 27 participants, and all analyses on BMI are therefore based on *n* = 11,125. Participants with valid FGF21 data were *n* = 880 (51% women).

**Table 1.**
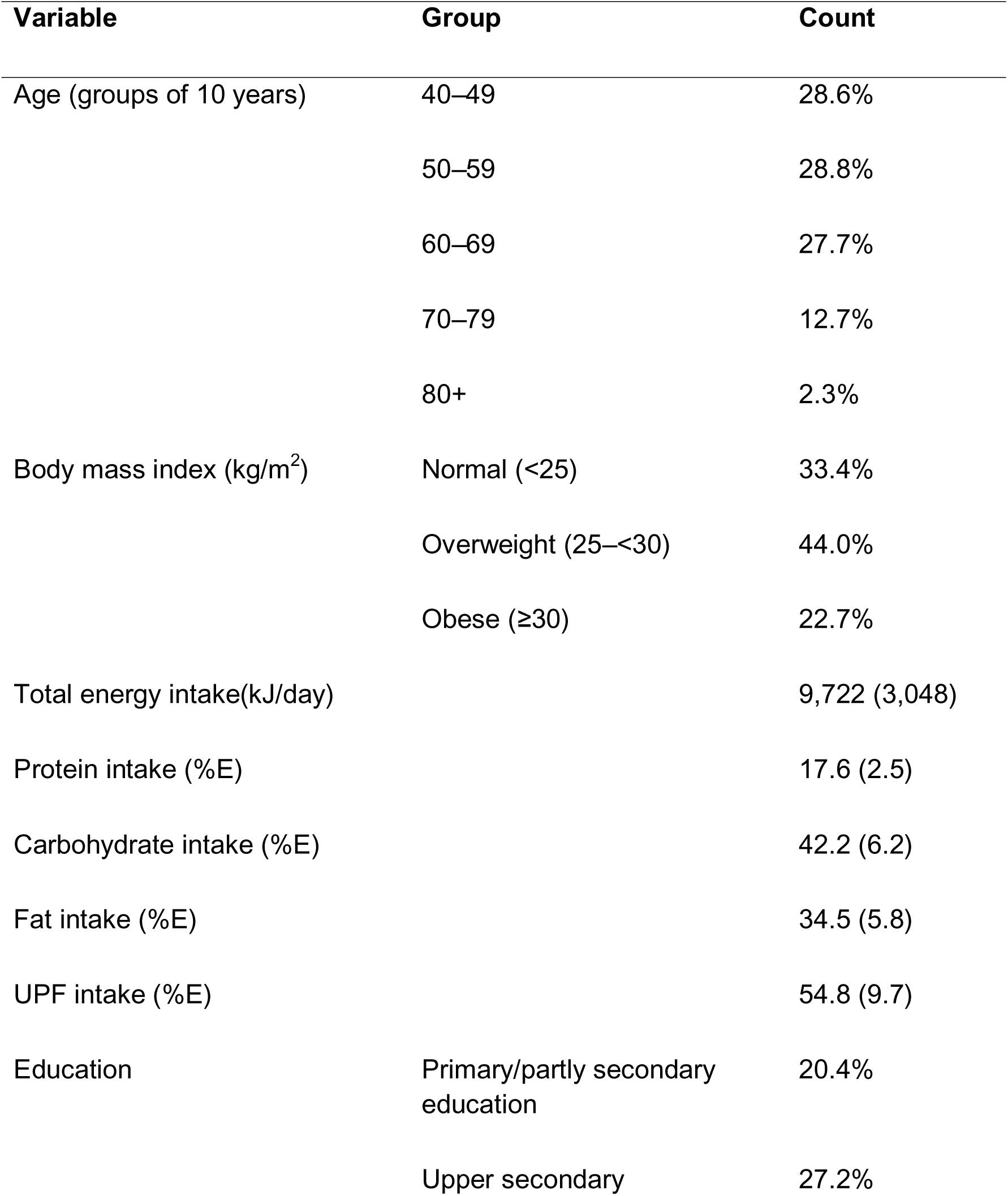
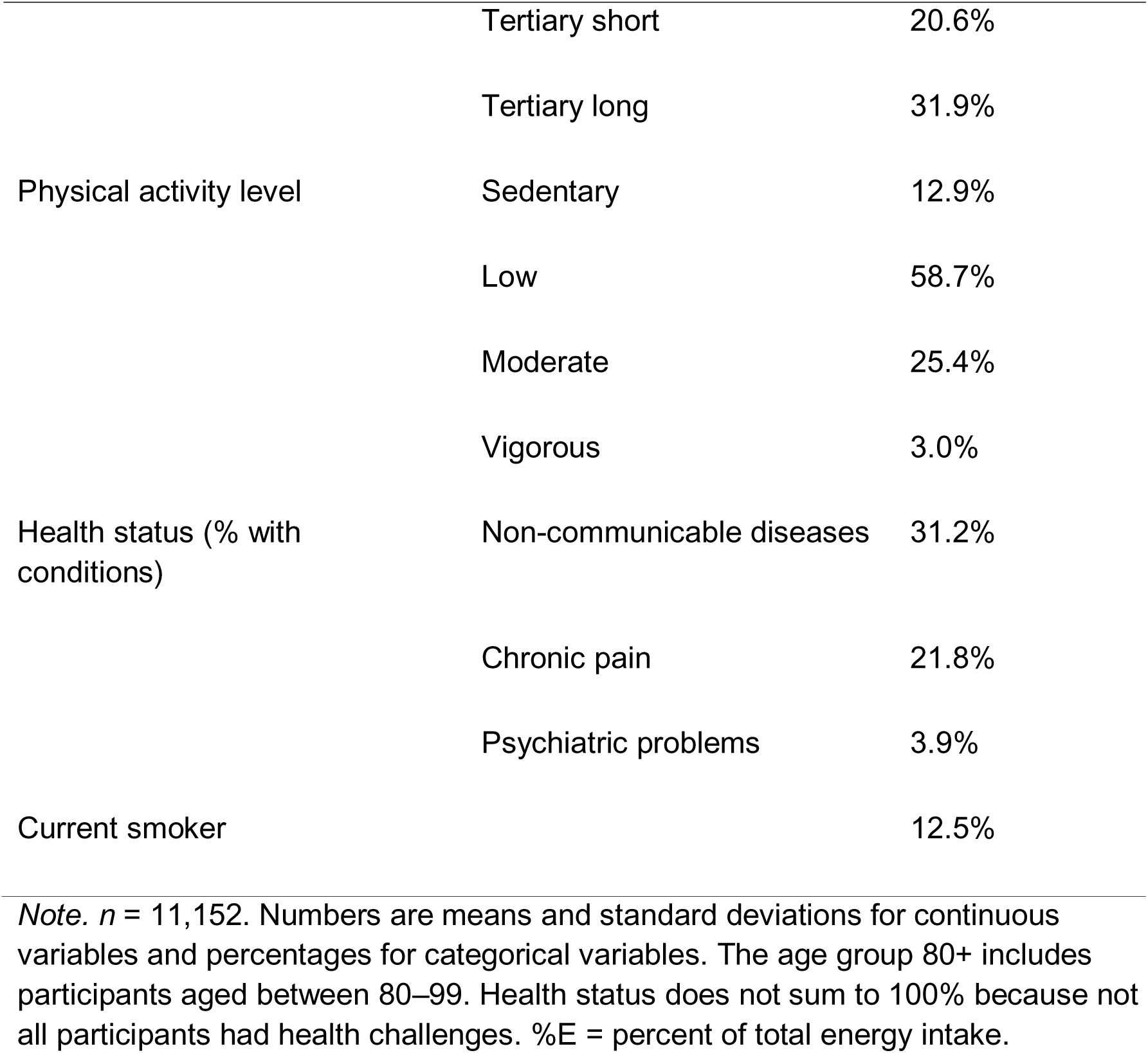
Descriptive statistics for the full sample from the Tromsø Study 2015–2016.

**Table 2.** Output from power functions with percent protein as predictor.

| Total energy intake<br>(kJ) |  | Energy density<br>(kJ/g) |  | Food dry weight (g) |  |
| --- | --- | --- | --- | --- | --- |
| *L* | *p* | *L* | *p* | *L* | *p* |
| -0.36 | < .001 | - .48 | < .001 | -0.31 | < .001 |

*Note.* Energy density is kJ/ wet weight of food; food dry weight is dry weight of all energy yielding nutrients.

### 3.2 The Protein Leverage Mechanism

Analyses of the strength of leverage protein has over energy intake found support for a partial protein leverage effect with the analysis showing that *L* = -0.36 (*p* < .001).

The negative *L*-value indicates that protein was negatively associated with energy intake (Figure 2A), in line with our hypothesis. The power function was fitted for carbohydrate and fat and the analysis revealed *L*-values of ∼0 (*p* = .966) and 0.33 (*p* < .001), respectively. This indicates that the proportion of carbohydrates in the diet had no control over energy intake (Figure 2B), while the proportion of fat was positively associated with energy intake (Figure 2C).

**Figure 2.**
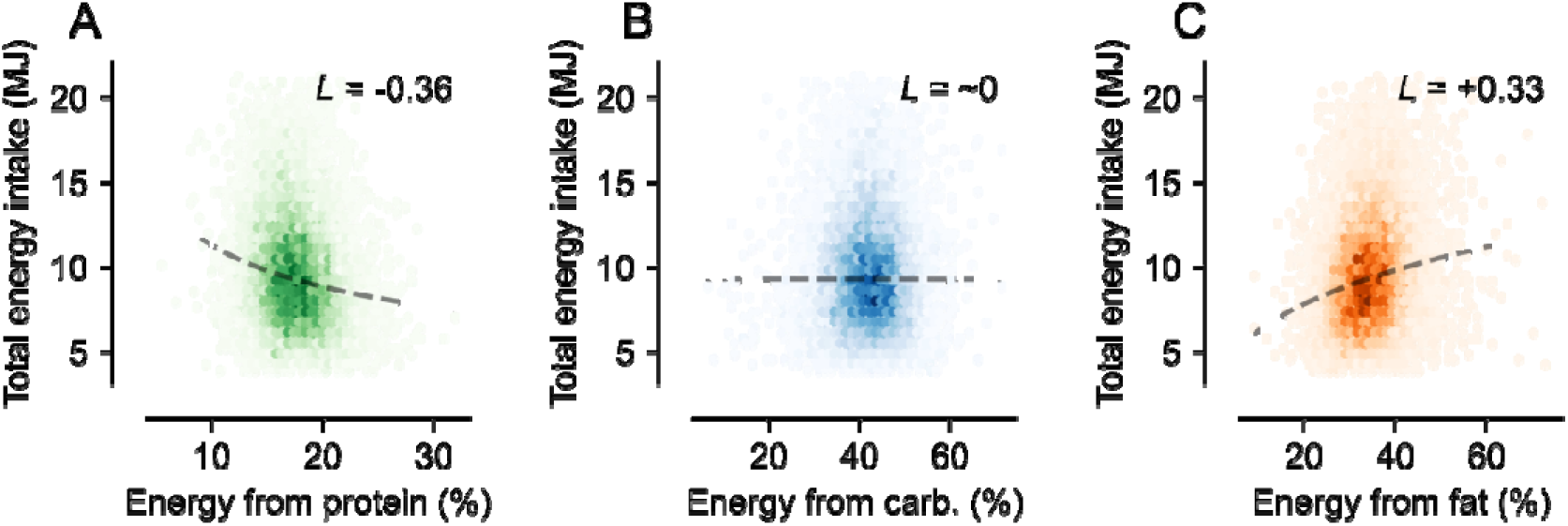
Power functions between total energy intake and proportional intake of each macronutrient. (A) shows protein, (B) shows carbohydrate, and (C) shows fat. Dashed lines show regression lines fitted with Equation 1.1. Data on individual participants are presented as a density heat map (darker colors indicate higher density of observations). *L* is the leverage strength of each macronutrient on energy intake where a positive number indicates that total energy intake increases as percentage of the macronutrient increases and a negative value indicates that total energy intake decreases as percentage of the macronutrient increases.

### 3.3 Mixture Models for Protein, Carbohydrate, and Fat

The AIC (Table S1) favored the second model (Table S2) which is visualized in the mixture triangle in Figure 3A which shows that the change in total energy intake mainly followed the protein axis, but fat also had an effect. As the proportion of protein increases from e.g., 10% to 30%, the predicted total energy intake decreases (as seen by the color change from yellow to purple). As the proportion of fat increases from e.g. 10% to 70%, the color changes from purple to orange-yellow.

**Figure 3.**
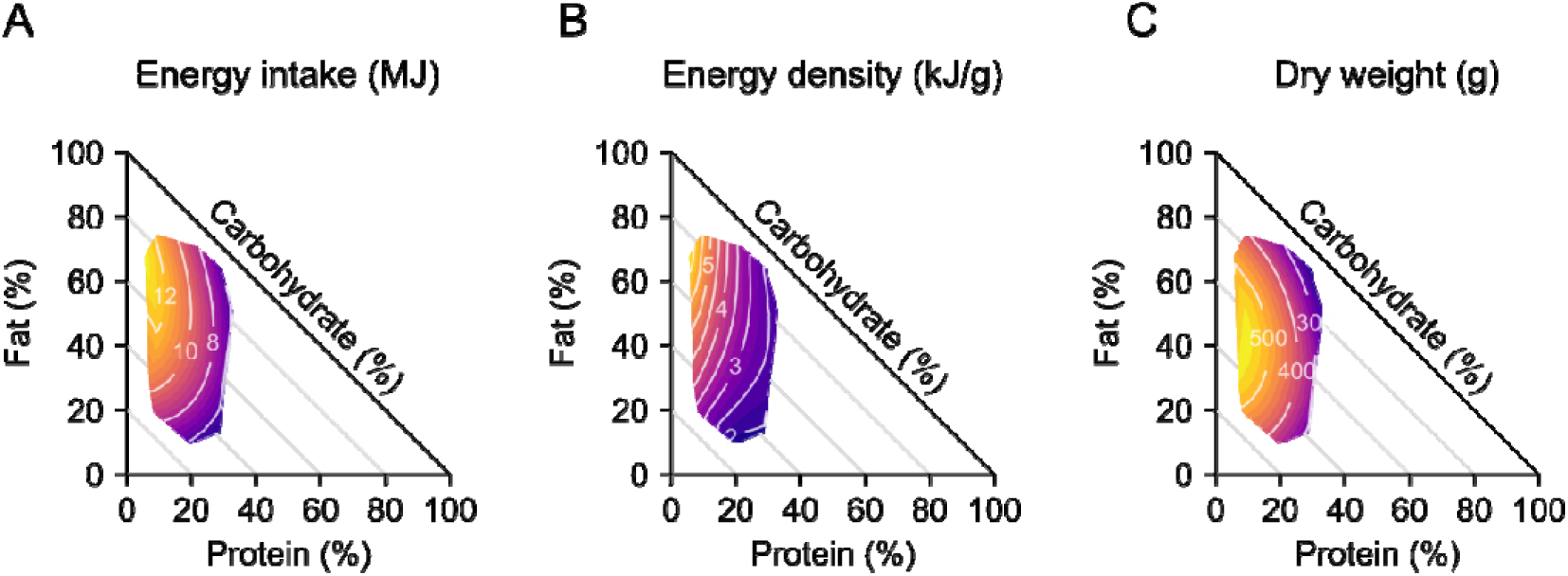
Equilateral mixture triangle for predicted total energy intake by macronutrient proportions. Predicted total energy intake based on the mixture model shown in Table S2 is shown as colored contours. Negatively sloped lines indicate percent protein, positively sloped lines indicate percent fat, and horizontal lines indicate percent carbohydrate. As percent protein increases, predicted energy intake decreases and as percent fat increases, predicted energy intake increases. Changes to percent carbohydrate have only a minor effect on predicted energy intake.

Additional analyses were run for energy density (Table S3) and dry weight of food (Table S4) to further probe macronutrient associations. Energy density (Figure 3B) is greatest in high fat/low protein diets, and lowest in high protein diets, regardless of the level of fat. Dry weight of food (Figure 3C) appears unaffected by fat but to be negatively associated with protein, and is thus highest in low protein diets, supporting a leveraging of protein on appetite.

### 3.4 Properties of Diets High in Ultra-Processed Foods

Several linear regression models were run to assess how changes in proportion of UPFs in the diet were related to (1) proportion of dietary protein, (2) dietary protein in absolute terms, and (3) total energy intake. The output from the regression models and estimated marginal mean comparisons are reported in the Supplementary Materials (Tables S5-S10). Proportion of UPFs in the diet was negatively related to proportion of dietary protein (Table S5) and positively related to total energy intake (Table S9), while the amount of protein in absolute terms stayed relatively stable between UPF quintiles (Table S7). Figure 4A summarizes these results.

**Figure 4.**
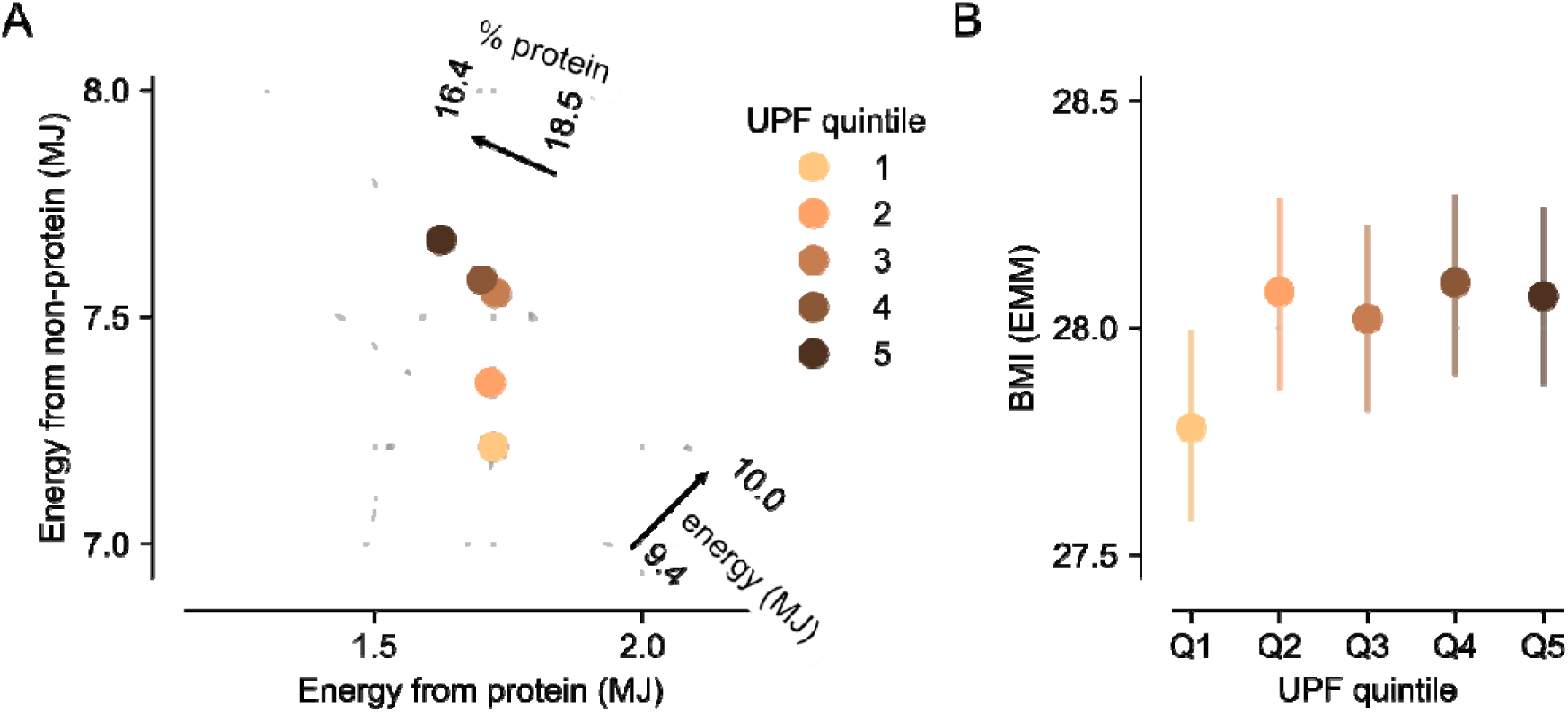
Relationship between ultra-processed foods (UPFs), dietary patterns, and BMI. (A) Descriptive bi-coordinate intake plot of protein versus non-protein energy intake (MJ) by quintiles (Qs) of UPF intake. Dots indicate mean intake of protein energy and non-protein energy (i.e., the combination of carbohydrate and fat) per quintile of UPF intake. The quintile dots most closely follow the protein prioritization line. Solid grey lines represent different predictions of intake for Q1 based on energy prioritization (negatively sloped line), protein prioritization (vertical line) or non-protein energy prioritization (horizontal line). Dashed grey lines show shift in total energy intake (negatively sloped) and protein intake (positively sloped) for Q5. *n* = 11,152. (B) Relationship between BMI and UPF intake plotted as quintiles. Data are estimated marginal means from regression model that control for sex, age, and physical activity. Error bars are 95%CI. *n* = 11,125.

Moreover, inspection of Figure 4A suggests that the leveraging effect of protein was greater for those consuming low levels of UPFs vs. those consuming high levels. As an exploratory analysis to test this, we performed a median split to divide participants into high and low UPF consumers and calculated the strength of protein leverage in each subpopulation (Figure 5A). Our analysis revealed that leverage was stronger in the low UPF group (*L* = -0.40, *p* < .001) than in the high UPF group (*L* = -0.29, p < .001). We confirmed that this difference was significant by shuffling labels which revealed the observed difference in L values to be greater than expected by chance (Figure 5B; p = 0.009).

**Figure 5.**
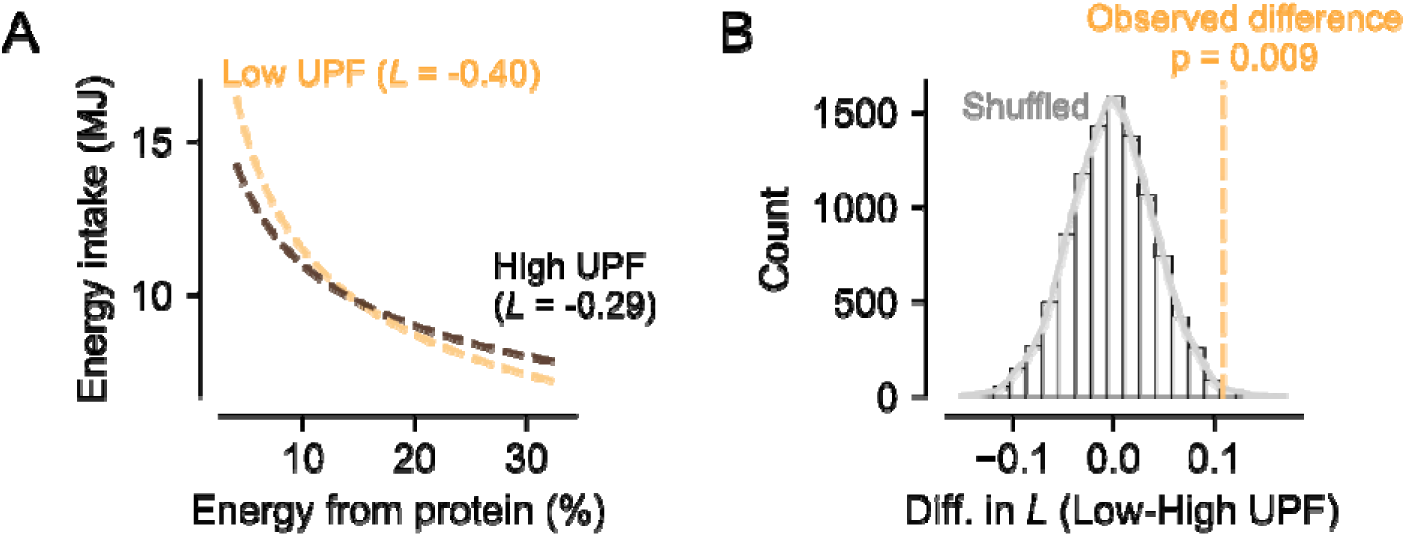
Protein leverage in low and high consumers of ultra-processed foods (UPFs). (A) Plotting of power function showing that the strength of leverage that protein exerts over total energy intake is greater in low vs. high consumers of UPFs after median split. (B) Histogram showing the distribution of the differences in *L* values between low and high UPF consumers after randomly shuffling labels 10,000 times. Dashed vertical line shows the observed difference from the real data.

### 3.5 Associations between Body Mass Index and Dietary Intake

The prediction that dietary protein is negatively associated with BMI builds on the assumption that energy intake will be negatively associated with BMI, we therefore tested this. We ran linear regression models of increasing complexity with covariates described under Demographics and Health Related Information and used AIC to choose among models. The AIC favored the model presented in the Supplementary Materials (Table S11), which revealed a weak and negative relationship between total energy intake and BMI (Table S12). Potential reasons for this lack of association are given in the Discussion. The lack of relationship means that these data are not suitable for testing associations between mechanisms mediating increased energy intake (i.e., protein leverage) and BMI. As the link between protein leverage and BMI was one of our pre-registered hypotheses we have nevertheless placed this analysis in Supplementary Materials (Table S14), however, results should be treated with caution.

In the analysis of the relationship between proportion of UPFs and BMI, the AIC (Table S15) favored the model that included UPF quintiles as well as the covariates age, sex, and physical activity level (see Table S16). The estimated marginal means were calculated for the quintiles of UPF intake while keeping the covariates in the model constant as the reference category (i.e., age = under 60 years, sex = female, physical activity level = sedentary) and while adjusting the *p*-value using the Dunnett method. This yielded a trend towards higher UPF quintiles being associated with a higher BMI as compared to the first UPF quintile (Table S17). The estimated marginal means per quintile of UPF are visualized in Figure 4B.

### 3.6 FGF21 and Dietary Protein

To assess the relationship between FGF21 and dietary protein, several multiple linear regression models were run, and the AIC-favored (Table S18) model is presented in Table 3 (*R^2^* = .04, *F*(7, 757) = 5.9, *p* < .001). The model reveals a negative, significant, relationship between dietary protein and FGF21, in line with our expectations. Several other covariates were also significantly associated with FGF21 including age, physical activity level, disease and smoking status.

**Table 3.** Regression model for fibroblast growth factor 21 by dietary protein.

| **Variable** | ***b*** | **SE** | ***t*** | ***p*** |
| --- | --- | --- | --- | --- |
| Intercept | 5.30 | 0.35 | 15.2 | < .001 |
| Protein, %E | -0.05 | 0.02 | -3.2 | .001 |
| Age | 0.28 | 0.09 | 3.0 | .002 |
| Sex | -0.12 | 0.08 | -1.4 | .168 |
| Physical activity | -0.28 | 0.13 | -2.1 | .035 |
| Disease | 0.20 | 0.08 | 2.4 | .016 |
| Education level | -0.02 | 0.098 | -0.2 | .848 |
| Smoking status | 0.34 | 0.13 | 2.7 | .008 |

*Note.* $n = 880$ . Covariates were dichotomous and coded as follows: age = 0 (under 60 years), sex = 0 (female), physical activity level = 0 (sedentary), disease = 0 (no disease), education level 0 ( $\leq 10$ years).

## 4. Discussion

In this study of 11,152 women and men from a general population in Norway, we examined predictions of the protein leverage hypothesis. We found strong support for the protein leverage mechanism (i.e., the energy intake regulating effects of dietary protein; *L* = -.36) in line with previous studies in the field (Honfo et al., 2023; Saner et al., 2023; Saner et al., 2020; Zhang et al., 2023). We did not find the predicted negative relationship between dietary protein and BMI, but given that there was no positive association between energy intake and BMI in our study, this was not surprising, as is discussed in more detail below. Our analyses of the relationship between dietary protein, total energy intake, and habitual UPF intake revealed relationships that are predicted to arise from the protein leverage hypothesis, namely, that a higher UPF intake was associated with greater energy intake but lower proportion of dietary protein. But again, our analysis failed to establish a concordant effect of UPF intake on BMI. Finally, in an exploratory analysis, we examined the relationship between FGF21 and dietary protein and this revealed the expected negative relationship.

### Positive leveraging effect of fat on energy intake

Our power functions not only revealed a partial protein leverage mechanism, but also a positive L-value for fat, again in line with previous studies (Honfo et al., 2023; Saner et al., 2023). The association between fat and energy intake may be related to several factors. A key consideration is that is that as protein leverages food intake, increases in fat will have a disproportionate effect on energy intake due to fat being calorically dense. This effect will be exacerbated in places where habitual diets are relatively high in fat. In Norway, as well as many other Western countries, fat makes up >30% of energy intake. In addition, other properties of fat may directly or indirectly augment this effect. For example, fat has a lower satiating effect than the other two macronutrients (Blundell & Macdiarmid, 1997), which could lead to larger meals and thus an excess energy intake when consuming diets high in fat. The high palatability of fat-rich foods (Mizushige et al., 2007) may also play a role in driving energy intake. As such, people may eat more when consuming diets high in fat because they taste better, or being exposed to high fat diets may lead to changes in dietary patterns such as larger meals or more snacking. The protein leverage hypothesis is based on a nutritional geometric framework and emphasizes that it is the balance of the macronutrients that are of importance for energy intake and potential weight gain and our results for fat and protein attest to this.

### Energy intake, protein and their association with BMI

Our second, key pre-registered hypothesis was that protein intake would be negatively correlated with BMI. This hypothesis rests on an assumption that protein leverages energy intake (as shown by hypothesis 1) and that this increased energy intake in turn leads to increased BMI. In our data, however, we failed to find support for this assumption, namely the relationship between total energy intake and BMI. The implication is that finding a link between protein and BMI would be highly unlikely – and indeed we did not find this. What then are the reasons for energy intake not positively predicting BMI in this sample? Importantly, there are several limitations when using self-reported food intake (e.g., Dhurandhar et al., 2015; Prentice et al., 2025), even when using a validated FFQ. Perhaps most significant, are the inherent difficulties in relating short-term dietary intake (as the FFQ measures) to long-term consequences such as BMI. This may be especially the case when examining a middle-aged to older sample, as in the present study. Weight gain resulting in increased BMI accumulates over a lifetime while the FFQ is designed to measure habitual food intake in a short, recent period. Moreover, people living with a higher BMI have been found to underreport energy intake to a larger degree, on a group level, than people in the normal BMI range (Howes et al., 2024; Poslusna et al., 2009). As a large proportion of the participants in our study are either living with overweight or obesity, this could have impacted our data. Indeed, calculations based on the Goldberg method (Black, 2000 as cited by; Lundblad et al., 2019) indicate that about 20% of the people in the Tromsø7 sample underreported energy intake when filling out the FFQ (Lundblad et al., 2019).

### Relationship between FGF21, energy expenditure and BMI

Ours and several other studies fail to find that low protein diets are related to a higher BMI, a key relationship predicted by the protein leverage mechanism (Honfo et al., 2023; Saner et al., 2023; Saner et al., 2020; Zhang et al., 2023). The role of the liver-derived hormone FGF21 in energy balance illustrates some important considerations in interpreting interactions between energy intake, macronutrient (im)balance, and resulting weight change. Here, we found that dietary protein was negatively associated with FGF21, an expected finding in line with results in prior studies (Gosby et al., 2016; Nicolaisen et al., 2025). Moreover, FGF21 is also suggested to upregulate energy expenditure (Nicolaisen et al., 2025). This has been shown most robustly in rodent models (Hill et al., 2019; Laeger et al., 2014) although one study has established this association in humans (Vinales et al., 2019). As such, the effect of low protein diets on energy balance may seem contradictory at first (protein leverage increases intake while inducing FGF21 which increases energy expenditure). However, these two effects can be tightly linked due to the established effect of surplus energy intake (induced by leverage) on triggering compensatory increases in expenditure. Thus, a key question - not just for protein leverage but for all mechanisms of obesity - is to determine the extent to which compensatory expenditure is able to offset increased intake. Moreover, what our the conditions or parameters that lead to this compensation being inadequate such that weight gain and obesity result from excess energy intake.

### Potential role of ultra-processed foods

A higher proportion of energy from UPFs was accompanied by a higher energy intake and a lower proportional protein intake. As detailed in the Results, a change in percent dietary protein was observed between the highest (16.4%) and lowest (18.5%) quintiles. This reduction by a mere 2% may seem modest, but this may have large effects on energy intake via leveraging. A previous RCT found that dilution by 3.5% of protein for a short period (14 days) may have contributed to significant increases in energy intake and subsequent weight gain in participants (Hall et al., 2019). Replotting of these RCT data by Raubenheimer and Simpson (2023) suggests that in this controlled setting, protein leverage could explain almost all increased energy intake associated with a UPF diet.

Subtle differences in dietary protein at a population-level have been associated with obesity. For example, by analyzing food balance sheets and available epidemiology, it was estimated that dilution of dietary protein coupled to perfect protein leverage could explain as much as two-thirds of observed weight gain in US adult population since the 1970s (Hall, 2019). In the current study, the *L*-value for protein was -0.36, which despite being less than perfect leverage, still means that protein leverage is a significant potential mechanism.

Whether the low protein nature of many UPFs is what leads these diets to cause excess energy intake is still under debate. Importantly, several other UPF properties could interact with protein content to increase energy intake. For example, UPF formulations, including the combination of macronutrients, encourage consummatory patterns that lead to high energy intake (Fazzino et al., 2019). In particular, high fat content may increase energy intake, independent of carbohydrate, given the positive *L*-value for fat found in our earlier analysis. Another possibility is that the umami and salt flavors associated with many UPFs may drive energy intake up (Grech et al., 2022b) due to the protein decoy effect. Foods rich in protein often have these savory flavors, and people in search of protein may use these cues to guide them towards a potentially nutritious food source. Dietary fiber (and energy density of food, which is inversely proportional), is low in UPFs, influences energy intake, and can reduce the protein leverage effect by limiting intake. Finally, UPFs are designed to increase food intake due to their industrialized, cosmetic, ingredients (Monteiro et al., 2019). All these factors may play a role in increasing energy intake in diets high in UPF content, in addition to the diluted protein content.

### Strengths and limitations

A major strength of the current study is the large sample size from a general population, high attendance, and data collection using standard methods and validated measurements. Another strength is that several of the findings replicate other studies, thus further strengthening the implications of the protein leverage hypothesis. Major limitations, as discussed above, are issues relating to self-reported food intake including potential underreporting and the difficulties in associating short-term estimation of food intake with long-term health outcomes, such as obesity.

## Conclusion

We found strong support for the protein leverage mechanism through several analysis methods, i.e., fitted power functions and mixture modeling. An interesting and related finding was the positive effect of proportion of fat on energy intake. Our study revealed that a diet consisting of a high proportion of UPFs was related to heightened energy intake and a lowered proportion of dietary protein, while protein intake in absolute terms remained relatively stable between UPF diets. Finally, we did not find a positive association between energy intake and BMI meaning that we were not able to properly test the predicted consequence of protein leverage increasing BMI.

## Data Availability

The data that support the findings of this study are available upon application to the Tromsoe Study. Restrictions apply to the availability of these data, which were used under license for this study.

## Author contributions

**Rikke Eriksen:** Conceptualization, Formal Analysis, Writing - Original Draft; **Kamilla Rognmo:** Conceptualization, Writing – Review and Editing; **Laila A Hopstock:** Conceptualization, Writing – Review and Editing, Investigation; **James E McCutcheon:** Conceptualization, Writing – Review and Editing, Visualization.

## Acknowledgements

The authors would like to acknowledge the staff involved in data collection and administration of the Tromsø Study and the participants that volunteered to take part. We would also like to acknowledge the manuscript’s reviewers for insightful and helpful comments during the review process. Funding was provided by Tromsø Research Foundation to JEM (19-SG-JMcC). The data that support the findings of this study are available upon application to the Tromsø Study. Restrictions apply to the availability of these data, which were used under license for this study.

