## Supplementary materials for "Evidence for a protein leverage effect on food intake in a Norwegian population"

### Supplementary Material

#### 1. NOVA Classification of food processing level

Group 1 includes unprocessed or minimally processed foods, with typical whole foods such as fruits, vegetables, meat, milk, and nuts. Group 2 includes ingredients that have been extracted from whole foods. They are often referred to as culinary ingredients, and contain ingredients such as oils, flours, and sugars. Group 3 includes processed foods, meaning foods that contain ingredients from Groups 1 and 2. It includes foods such as homemade bread, cheese, and smoked fish (Huybrechts et al., 2022). Group 4 is the UPF group. Typical products are industrial bread, breakfast cereals, and chips (Monteiro, 2009). These foods are often processed in a way or contain ingredients that cannot be found in a private kitchen and so has been made industrially.

The procedure started with a single rater categorizing all items in the FFQ. Foods belonging to NOVA groups one and two are usually single ingredient items, and thus unambiguously belong to a single food group. Foods in groups three and four consist of several ingredients and their categorization is thus more complex. To account for this, FFQ items consisting of more than one ingredient were scored as a lower-bound and upper-bound scenario. The lower bound scenario was the least processed version of the item, e.g., home-made lasagna, while the upper-bound scenario was the most processed version, e.g., ready-to-heat, store bought lasagna. The home-made dishes were decomposed into their ingredients based on Norwegian recipes and the ingredients were classified into NOVA groups. This ingredient list was then the basis for categorizing the lower-bound scenario. Categorization of store-bought products was based on ingredient lists per item. Any items with ingredients from the list in Appendix D were categorized as belonging to Group 4 (UPFs). This list was adopted from Huybrechts et al. (2022) as based on Monteiro (2009). This categorization procedure resulted in an spreadsheet with a list of the FFQ items, and their corresponding lower- and upper-bound scenario. This was the basis for further categorization.

The list of FFQ items and the scenarios were then used by two raters (one of them is the initial rater of the upper-and lower-bound scenarios) to select the middle-bound scenario per item. The middle-bound scenario was the most likely scenario of the lower-and upper-bound-scenarios based on the sample characteristics (Tromsø inhabitants aged > 40 years old in 2015-2016). The ratings by the two raters of the middle-bound scenarios were assessed by an inter-rater reliability test (reported in Appendix E), which indicated moderate agreement between raters. Therefore, a list of items scored differently by the two raters (*n* = 23) (see Appendix F) was scored by an independent set of raters (*n* = 5) to find the most likely scenario. This group of raters were all over the age of 40 and were living in Tromsø at the time of the data collection (2015-2016). They were instructed to consider local habits in their judgement. The median score for each item was used as the middle-bound scenario and this list was used to make a binary variable indicating whether an item was ultra-processed or not.

#### 2. Mixture modeling of the relationship between macronutrients and total energy intake

Mixture model analysis was carried out in accordance with (Lawson & Willden, 2016). The first four Scheffé models were ran on the predictors.

The first model is a linear model written by Scheffé (Lawson & Willden, 2016; Scheffé, 1958) as:

$$y=\sum_{i=1}^{q} \beta_{i}x_{i}+\epsilon$$

The second model (quadratic):

$$y=\sum_{i=1}^{q} \beta_{i}x_{i}+ \sum_{i=1}^{q-1} \sum_{j=i+1}^{q} \beta_{ij}x_{i}x_{j}+\epsilon$$

The third model (full cubic):

$$y=\sum_{i=1}^{q} \beta_{i}x_{i}+ \sum_{i=1}^{q-1} \sum_{j=i+1}^{q} \beta_{ij}x_{i}x_{j}+ \sum_{i=1}^{q-1} \sum_{j=i+1}^{q} \delta_{ij}x_{i}x_{j}(x_{i}{+x}_{j})+\sum_{i=1}^{q-2} \sum_{j=i+1}^{q-1} \sum_{k=j+1}^{q} \beta_{ijk}x_{i}x_{j}x_{k}+\epsilon$$

The fourth model (special-cubic):

$$y=\sum_{i=1}^{q} \beta_{i}x_{i}+ \sum_{i=1}^{q-1} \sum_{j=i+1}^{q} \beta_{ij}x_{i}x_{j}+\sum_{i=1}^{q-2} \sum_{j=i+1}^{q-1} \sum_{k=j+1}^{q} \beta_{ijk}x_{i}x_{j}x_{k}+\epsilon$$

Models 2, 3 and 4 were within 2 AIC points from each other, and so the simplest model were chosen, i.e., model 2.

#### 3. Supplementary statistics

**Table S1 |** AIC Comparison for Regression Models for the Four Scheffé Polynomials

|  | k | AIC | ∆AIC | AIC weight | Log-likelihood |
| --- | --- | --- | --- | --- | --- |
| Model 0 | 2 | 210582 | 684 | 0 | -105289 |
| Model 1 | 4 | 209939 | 42 | 0 | -104966 |
| Model 2 | 7 | 209899 | 1.10 | .25 | -104942 |
| Model 3 | 11 | 209899 | 0.71 | .31 | -104938 |
| Model 4 | 8 | 209898 | 0 | .44 | -104941 |

Note. AIC favored model shown in bold. If several models were within two AIC points of each other, the simplest model would be chosen.

**Table S2 |** Mixture Model for Total Energy Intake

| Variable | b | SE | t | p |
| --- | --- | --- | --- | --- |
| Protein | -30,508 | 15,755 | -1.94 | .053 |
| Carbohydrate | 3,525 | 1,442 | 2.45 | .015 |
| Fat | 10,924 | 2,213 | 4.94 | <.001 |
| Protein * fat | 31,457 | 26,423 | 1.19 | .234 |
| Protein * carbohydrate | 43,152 | 23,525 | 1.83 | .067 |
| Fat* carbohydrate | 25,751 | 4,389 | 5.87 | <.001 |

Note. n = 11,152 from Tromsø7. %E = percent of total energy intake from respective macronutrient. SE = standard error.

**Table S3 |** Mixture Model for Energy Density

| Variable | b | SE | t | p |
| --- | --- | --- | --- | --- |
| Protein | 15.1 | 4.30 | 3.5 | <.001 |
| Carbohydrate | 8.4 | 0.60 | 13.9 | <.001 |
| Fat | 2.3 | 0.39 | 5.9 | <.001 |
| Protein * fat | -39.0 | 7.21 | -5.4 | <.001 |
| Protein * carbohydrate | -22.6 | 6.42 | -3.5 | <.001 |
| Fat* carbohydrate | 5.3 | 1.20 | 4.4 | <.001 |

Note. n = 11,152 from Tromsø7. %E = percent of total energy intake from respective macronutrient. SE = standard error.

**Table S4 |** Mixture Model for Intake of Dry Weight of Food

| Variable | b | SE | t | p |
| --- | --- | --- | --- | --- |
| Protein | -1804.8 | 759.7 | -2.38 | .018 |
| Carbohydrate | 188.6 | 106.7 | 1.77 | .072 |
| Fat | 322.3 | 69.5 | 4.64 | <.001 |
| Protein * fat | 2739.8 | 1274.1 | 2.15 | .032 |
| Protein * carbohydrate | 2357.2 | 1134.4 | 2.08 | .038 |
| Fat* carbohydrate | 1331.4 | 211.7 | 6.29 | <.001 |

Note. n = 11,152 from Tromsø7. %E = percent of total energy intake from respective macronutrient. SE = standard error.

**Table S5 |** Regression Model for Percent Protein by Quintiles of Ultra-Processed Food

| Variable | b | SE | t | p |
| --- | --- | --- | --- | --- |
| Intercept | 18.45 | 0.05 | 357.3 | < .001 |
| Quintile 2 | -0.41 | 0.07 | -5.6 | < .001 |
| Quintile 3 | -0.72 | 0.07 | -9.9 | < .001 |
| Quintile 4 | -1.11 | 0.07 | -15.1 | < .001 |
| Quintile 5 | -2.01 | 0.07 | -27.5 | < .001 |

Note. n = 11,152. Quintiles represent the proportion of ultra-processed foods (UPFs) as a percentage of total food intake (%E) in the diet. The first quintile is handled in the intercept and used as the reference point for comparison in the regression model. R2 = .07, F(4, 11147) = 219, p < .001.

**Table S6 |** Estimated mean differences between Quintile 1 and other quintiles of dietary protein (%) by ultra-processed food intake

| Contrast | Estimate | SE | t ratio | p |
| --- | --- | --- | --- | --- |
| Quintile 2- Quintile 1 | -0.41 | 0.073 | -5.599 | < .001 |
| Quintile 3- Quintile 1 | -0.72 | 0.073 | -9.897 | < .001 |
| Quintile 4- Quintile 1 | -1.11 | 0.073 | -15.140 | < .001 |
| Quintile 5- Quintile 1 | -2.0 | 0.073 | -27.468 | < .001 |

Note. n = 11,152. Quintiles represent the proportion of ultra-processed foods (UPFs) as a percentage of total food intake (%E) in the diet. Comparisons are between the first and all higher quintiles of UPF intake. The p-value was corrected for multiple comparisons using the Dunnett method.

**Table S7 |** Regression Model for Protein in Absolute Terms by Quintiles of Ultra-Processed Food

| Variable | b | SE | t | p |
| --- | --- | --- | --- | --- |
| Intercept | 101.30 | 0.68 | 148.34 | < .001 |
| Quintile 2 | -0.34 | 0.97 | -0.35 | .724 |
| Quintile 3 | 0.28 | 0.97 | 0.29 | .774 |
| Quintile 4 | -1.24 | 0.97 | -1.28 | .199 |
| Quintile 5 | -5.70 | 0.97 | -5.90 | < .001 |

*Note. n* = 11,152. Quintiles represent the proportion of ultra-processed foods (UPFs) as a percentage of total food intake (%E) in the diet. The first quintile is handled in the intercept and used as the reference point for comparison in the regression model. *R^2^* = .004, *F*(4, 11147) = 13, *p* < .001

**Table S8 |** Estimated mean differences between Quintile 1 and other quintiles of dietary protein (g) by ultra-processed food intake

| Contrast | Estimate | SE | t ratio | p |
| --- | --- | --- | --- | --- |
| Quintile 2- Quintile 1 | -0.34 | 0.97 | -0.35 | .970 |
| Quintile 3- Quintile 1 | 0.28 | 0.97 | 0.29 | .981 |
| Quintile 4- Quintile 1 | -1.24 | 0.97 | -1.28 | .501 |
| Quintile 5- Quintile 1 | -5.70 | 0.97 | -5.90 | <.0001 |

Note. n = 11,152. Quintiles represent the proportion of ultra-processed foods (UPFs) as a percentage of total food intake (%E) in the diet. Comparisons are between the first and all higher quintiles of UPF intake. The p-value was corrected for multiple comparisons using the Dunnett method.

**Table S9 |** Regression Model for Total Energy Intake by Quintiles of Ultra-Processed Food

| Variable | b | SE | t | p |
| --- | --- | --- | --- | --- |
| Intercept | 9408 | 64 | 146.1 | < .001 |
| Quintile 2 | 172 | 91 | 1.9 | .060 |
| Quintile 3 | 410 | 91 | 4.5 | < .001 |
| Quintile 4 | 437 | 91 | 4.8 | < .001 |
| Quintile 5 | 554 | 91 | 6.1 | < .001 |

*Note. n* = 11,152. Quintiles represent the proportion of ultra-processed foods (UPFs) as a percentage of total food intake (%E) in the diet. The first quintile is handled in the intercept and used as the reference point for comparison in the regression model. *R^2^* = .004, *F*(4, 11147) = 12, *p* < .001

**Table S10 |** Estimated mean differences between Quintile 1 and other quintiles of ultra-processed food intake by total energy intake

| Contrast | Estimate | SE | t ratio | p |
| --- | --- | --- | --- | --- |
| Quintile 2- Quintile 1 | 172 | 91.1 | 1.9 | .188 |
| Quintile 3- Quintile 1 | 410 | 91.1 | 4.5 | <.0001 |
| Quintile 4- Quintile 1 | 437 | 91.1 | 4.8 | <.0001 |
| Quintile 5- Quintile 1 | 554 | 91.1 | 6.1 | <.0001 |

Note. n = 11,152. Quintiles represent the proportion of ultra-processed foods (UPFs) as a percentage of total food intake (%E) in the diet. Comparisons are between the first and all higher quintiles of UPF intake. The p-value was corrected for multiple comparisons using the Dunnett method.

**Table S11 |** AIC Comparison for Regression Models for the Relationship Between BMI and Total Energy Intake

|  | Variable | k | AIC | ∆AIC | AIC weight | Log-likelihood |
| --- | --- | --- | --- | --- | --- | --- |
| Model 1 | Protein, %E | 3 | -10043 | 267.0 | 0 | 5025 |
| Model 2 | + age | 4 | -10048 | 262.7 | 0 | 5028 |
| Model 3 | + sex | 5 | -10223 | 81 | 0 | 5117 |
| Model 4 | + physical activity | 6 | -10310 | 0.0 | 1 | 5161 |
| Model 5 | + health variables | 7 | -9746 | 564.8 | 0 | 4880 |
| Model 6 | + education | 8 | -9691 | 619.4 | 0 | 4853 |
| Model 7 | + smoke | 9 | -9728 | 582.2 | 0 | 4873 |

*Note. n* = 11,125. The AIC favored model is shown in bold. %E = percent of total energy intake from protein.

**Table S12 |** Regression model for log-transformed body mass index by total energy intake

| Variable | b | SE | t | p |
| --- | --- | --- | --- | --- |
| Intercept | 3.347 | 0.006 | 534 | < .001 |
| Energy kJ | -0.000001 | 0.000 | -2.26 | 0.024 |
| Age | 0.0037 | 0.003 | 1.25 | 0.212 |
| Sex | 0.0040 | 0.003 | 13.22 | < .001 |
| Physical activity | 0.0774 | 0.004 | -17.90 | < .001 |

*Note. n* = 11,125. Covariates were coded as follows: age = 0 (under 60 years), sex = 0 (female), physical activity level = 0 (sedentary).

In the analyses of the relationship between protein intake and BMI, the AIC (Table S11) favored the model presented in Table S12, which included age, sex, and physical activity (*R^2^* = .07, *F*(4, 10866) = 204, *p* < .001). The model indicated a positive relationship between protein intake and BMI, a finding contrary to our hypothesis. The covariates sex and physical activity level were also significant in the model. This shows that being male was associated with a higher BMI than being female and being physically active was associated with a lower BMI than being sedentary. Age was not significant in the model, but to check for interaction effects, a follow-up analysis was run and reported in the Table S13. The main effect of age was not significant, but the interaction between dietary protein and age was significant and positive. This interaction is visualized in Figure S1.

**Table S13 |** AIC Comparison for Regression Models for the Relationship Between BMI and Dietary Protein

|  | Variable | k | AIC | ∆AIC | AIC weight | Log-likelihood |
| --- | --- | --- | --- | --- | --- | --- |
| Model 1 | Energy kJ | 3 | -10,260 | 332 | 0 | 5,133 |
| Model 2 | + age | 4 | -10,260 | 332 | 0 | 5,134 |
| Model 3 | + sex | 5 | -10,470 | 122 | 0 | 5,240 |
| Model 4 | + physical activity | 6 | **-10,592** | **0** | 1 | 5,302 |
| Model 5 | + health variables | 7 | -9,982 | 610 | 0 | 4,998 |
| Model 6 | + education | 8 | -9,916 | 675 | 0 | 4,966 |
| Model 7 | + smoke | 9 | -9,95 | 641 | 0 | 4,985 |

*Note. n* = 11,125. The AIC favored model is shown in bold.

**Table S14 |** Regression model for log-transformed body mass index by dietary protein

| Variable | b | SE | t | p |
| --- | --- | --- | --- | --- |
| Intercept | 3.20 | 0.011 | 293.0 | < .001 |
| Protein, %E | 0.01 | 0.001 | 17.0 | < .001 |
| Age | 0.01 | 0.003 | 0.5 | .638 |
| Sex | 0.04 | 0.003 | 14.8 | < .001 |
| Physical activity | -0.08 | 0.004 | -18.9 | < .001 |

*Note. n* = 11,125. Covariates were coded as follows: age = 0 (under 60 years), sex = 0 (female), physical activity level = 0 (sedentary). %E = percent of total energy intake from protein.

**Table S15 |** AIC Comparison for Regression Models for the Relationship Between Body Mass Index and Ultra-Processed Food Proportion

|  | Variable | k | AIC | ∆AIC | AIC weight | Log-likelihood |
| --- | --- | --- | --- | --- | --- | --- |
| Model 1 | Quintiles | 6 | -10,081 | 227 | 0 | 5,046 |
| Model 2 | + age | 7 | -10,088 | 219 | 0 | 5,051 |
| Model 3 | + sex | 8 | -10,224 | 84 | 0 | 5,120 |
| Model 4 | + physical activity | 9 | **-1**0308 | **0** | 1 | 5,163 |
| Model 5 | + health variables | 10 | -9,743 | 565 | 0 | 4,881 |
| Model 6 | + education | 11 | -9,688 | 619 | 0 | 4,855 |
| Model 7 | + smoke | 12 | -9,729 | 578 | 0 | 4,877 |

*Note. n* = 11,125. Quintiles represent the proportion of ultra-processed foods (UPFs) as a percentage of total food intake (%E) in the diet. The AIC favored model is shown in bold.

**Table S16 |** Regression model for log-transformed body mass index by UPF quintiles

| Variable | b | SE | t | p |
| --- | --- | --- | --- | --- |
| Intercept | 3.328 | 0.005 | 624.6 | < .001 |
| Quintile 2 | 0.011 | 0.005 | 2.3 | 0.0192 |
| Quintile 3 | 0.010 | 0.005 | 1.9 | 0.0531 |
| Quintile 4 | 0.012 | 0.005 | 2.5 | 0.0118 |
| Quintile 5 | 0.011 | 0.005 | 2.2 | 0.0256 |
| Age | 0.005 | 0.003 | 1.8 | 0.0744 |
| Sex | 0.037 | 0.003 | 12.2 | < .001 |
| Physical activity | -0.077 | 0.004 | -17.9 | < .001 |

*Note. n* = 11,125. Quintiles represent the proportion of ultra-processed foods (UPFs) as a percentage of total energy intake (%E) in the diet. The first quintile is handled in the intercept and used as the reference point for comparison in the regression model. Covariates were dichotomous and coded as follows: age = 0 (under 60 years), sex = 0 (female), physical activity level = 0 (sedentary). (*R^2^* = .05, *F*(7, 10863) = 74, *p* < .001).

**Table S17 |** Estimated mean differences between Quintile 1 and other quintiles of BMI by ultra-processed food intake

| Contrast | Estimate | SE | t ratio | p |
| --- | --- | --- | --- | --- |
| Quintile 2- Quintile 1 | 0.011 | 0.005 | 2.3 | .067 |
| Quintile 3- Quintile 1 | 0.009 | 0.005 | 1.9 | .169 |
| Quintile 4- Quintile 1 | 0.012 | 0.005 | 2.5 | .042 |
| Quintile 5- Quintile 1 | 0.011 | 0.005 | 2.2 | .087 |

*Note. n* = 11,125. Quintiles represent the proportion of ultra-processed foods (UPFs) as a percentage of total food intake (%E) in the diet. Comparisons are between the first and all higher quintiles of UPF intake. Means are adjusted for covariates which were coded as follows: age = 0 (under 60 years), sex = 0 (female), physical activity level = 0 (sedentary). The p-value was corrected for multiple comparisons using the Dunnett method.

**Table S18 |** AIC Comparison for Regression Models for the Relationship Between Fibroblast Growth Factor 21 and Dietary Protein

|  | Variable | k | AIC | ∆AIC | AIC weight | Log-likelihood |
| --- | --- | --- | --- | --- | --- | --- |
| Model 1 | Protein, %E | 3 | 2712 | 352 | 0 | -1353 |
| Model 2 | + age | 4 | 2705 | 344 | 0 | -1348 |
| Model 3 | + sex | 5 | 2704 | 344 | 0 | -1347 |
| Model 4 | + physical activity | 6 | 2613 | 253 | 0 | -1301 |
| Model 5 | + health variables | 7 | 2386 | 25 | 0 | -1186 |
| Model 6 | + education | 8 | 2381 | 20 | 0 | -1182 |
| Model 7 | + smoke | 9 | 2360 | 0 | 1 | -1171 |

*Note. n* = 880. The AIC favored model is shown in bold. %E = percent of total energy intake from protein.
